# Conflicts of Interest of Pediatric Journal Authors

**DOI:** 10.1101/2021.05.22.21257647

**Authors:** Rebecca L Petlansky, Amadea D Bekoe-Tabiri, Vanessa N Bueno, AnnMarie N Onwuka, Michael R Gionfriddo, Brian J Piper

**Affiliations:** Geisinger Commonwealth School of Medicine, Scranton, PA 18510; Bryn Mawr College, Bryn Mawr, PA 19010; The University of Scranton, Scranton, PA 18510; Center for Pharmacy Innovation and Outcomes, Geisinger, Forty Fort PA 18704; School of Pharmacy, Duquesne University, Pittsburgh, PA 15282

## Abstract

The Physician Payments Sunshine Act requires disclosure of payments made by drug and medical device manufacturers to physicians or teaching hospitals. Academic literature extensively documents gender disparities in the medical profession with regard to salary, promotion, and government funded research. This investigation sought to quantify potential conflicts of interest (CoIs) in pediatric medical journals, specifically examining sex differences. To identify potential CoIs, we examined manuscripts published prior to 2019 in six pediatric journals (*JAMA Pediatrics, Pediatrics, The Journal of Pediatrics, Pediatric Blood and Cancer, Pediatric Critical Care Medicine*, and *The Pediatric Infectious Disease Journal*). We collected physician demographics and specialty from the National Plan and Provider Enumeration System National Provider Identifier Registry and compensation data from both ProPublica’s Dollars for Docs (PDD) and Centers for Medicare and Medicaid Services Open Payments (CMSOP). Data was collected on 2,747 authors from 929 manuscripts. Of the 1,088 authors based in the United States with medical degrees (40.5% female), 510 (46.9%) had entries in PDD and CMSOP. Overall, 11,791 payments to these physician-authors totaled $9,586,089.97. Males were 19.6% more likely to receive payments (RR = 1.20, 95% CI [1.05, 1.37], p = 0.008). The mean amount received by males was $23,250.71 while that received by females was $10,970.78 (mean difference = 12,279.92, 95% CI = (2036.31, 22,523.54), p = 0.019). A comprehensive understanding of these CoIs can inform the disclosure policies of journals to promote transparency of authors.

---

A conflict of interest (CoI) exists when judgment regarding an individual’s primary interest, such as patient welfare or research integrity, may be unduly influenced by secondary interests, such as financial interests (1). Medical journals require authors to disclose their relationships with pharmaceutical and medical device companies because such relationships can serve as potential CoIs. However, these potential CoIs are not always reported and disclosures are rarely verified (2,3).

Verification of CoIs, was facilitated by the passage of the Physician Payments Sunshine Act, which requires public reporting of payments made to physicians and teaching hospitals from pharmaceutical and medical device companies (4). This payment data can be explored through the use of databases such as the Centers for Medicare and Medicaid Services Open Payments (CMSOP) program and ProPublica’s Dollars for Docs (PDD) (5, 6). CMSOP is a systematic nationwide effort to report these payments to the public. PDD database also contains payment information made to physicians and teaching hospitals and serves as a tool to search these payments.

Several studies have characterized CoIs including studies examining guidelines (7), point of care databases (8), textbooks (9, 10), and journal articles (11). Additionally, several reports have documented a disparity in payments between males and females (12, 13, 14).

One area where CoIs have not been characterized is pediatrics. The proportion of women in pediatrics has grown over the past several decades such that women now represent a majority in pediatricians (15,16). Our objective was to quantify potential CoIs of authors publishing in pediatric medical journals and determine any differences in payments to male and female physicians.

## Methods

### Procedures

A total of six pediatric medical journals were selected based on journal rankings on Scimago Journal and Country Rank (17), a publicly available portal that provides bibliometric indicators of journals. We attempted to select high impact journals with a large proportion of authors who are physicians located in the United States. We chose three general pediatric journals (*JAMA Pediatrics* 2018 Scimago Journal Rank (SJR) = 5, *Pediatrics* SJR = 3, and *The Journal of Pediatrics* SJR = 1.2) and three pediatric subspecialty journals (*The Pediatric Infectious Disease Journal* SJR = 1.3, *Pediatric Blood and Cancer* SJR = 1.3, and *Pediatric Critical Care Medicine* SJR = 1.3).

Within each journal, we began examining articles published in December 2018 and continued in reverse chronological order with the intention to retrieve at least 100 authors with entries in CMSOP (3) and PDD (4).

For each article, the first author’s name was recorded. If the author had a medical degree and was located in the United States, their name was entered into both CMSOP and PDD to determine if they had received financial compensation from pharmaceutical or medical device companies.

CMSOP and PDD contain data from payment reports released by the Centers for Medicare and Medicaid Services. Physicians are included in the database if payments total $10 or more. CMSOP categorizes payments as general payments, research payments, associated research funding, and ownership and investment interest. General payments include fees related to consulting, services other than consulting, travel and lodging, food and beverage, honoraria, education, royalties or licenses, speaking at an accredited/certified education program, and speaking at an unaccredited/non-certified education program. A total payment amount and number of payments is provided. Payment amounts and number of payments provided by specific companies or within specific categories of payment can also be viewed. PDD also provides a total payment amount and number of payments in the category of general payments, which includes promotional speaking, consulting, meals, travel and royalties.

Financial compensation that was received by the authors between 2015-2018 (a timeframe that included 36 months prior to publication, the standard for reporting potential CoIs) and that was classified as ‘general payments’ on CMSOP was recorded. Payments classified as research payment, associated research funding, or ownership and investment interest were not included. Total payment amount and number of payments on PDD was also recorded for each of the years.

The author’s name was also entered into the National Plan and Provider Enumeration System National Provider Identifier (NPPES NPI) Registry and degree (DO or MD), sex, specialty, and state of practice were also recorded.

This process was repeated for the last author of the article as well as a middle author. Middle author was chosen based on their possession of a medical degree. If multiple middle authors had medical degrees, the individual with the highest compensation according to PDD and CMSOP was reported for inclusiveness.

### Data Analysis

We summarized general payments to physicians using descriptive statistics including proportion of physicians receiving payments and amounts received. We also conducted analyses based on journal type (general vs. specialty), sex (male vs. female), and payment database (CMSOP and PDD). Payment database was examined as prior work identified minor discrepancies across databases (9). To compare across groups, we utilized t-tests and Mann Whitney tests for continuous data and chi-square tests for categorical data. All statistical analysis was performed in SYSTAT (Version 13, Systat Software, Inc, San Jose, California). GraphPad Prism (Version 8, GraphPad Software Inc., San Diego, California) was used to generate figures.

## Results

We examined 2,747 authors from 929 manuscripts. Of these authors, 1,659 were excluded because they did not hold a medical degree or were located outside of the U.S. (Figure 1). Of 1,088 authors that were U.S. based physicians (40.5% female), 510 (36.3% female) had received general payments within 2015 to 2018. Overall, 11,791 payments to these 510 physicians totaled $9,586,089.97.

**Figure 1.**
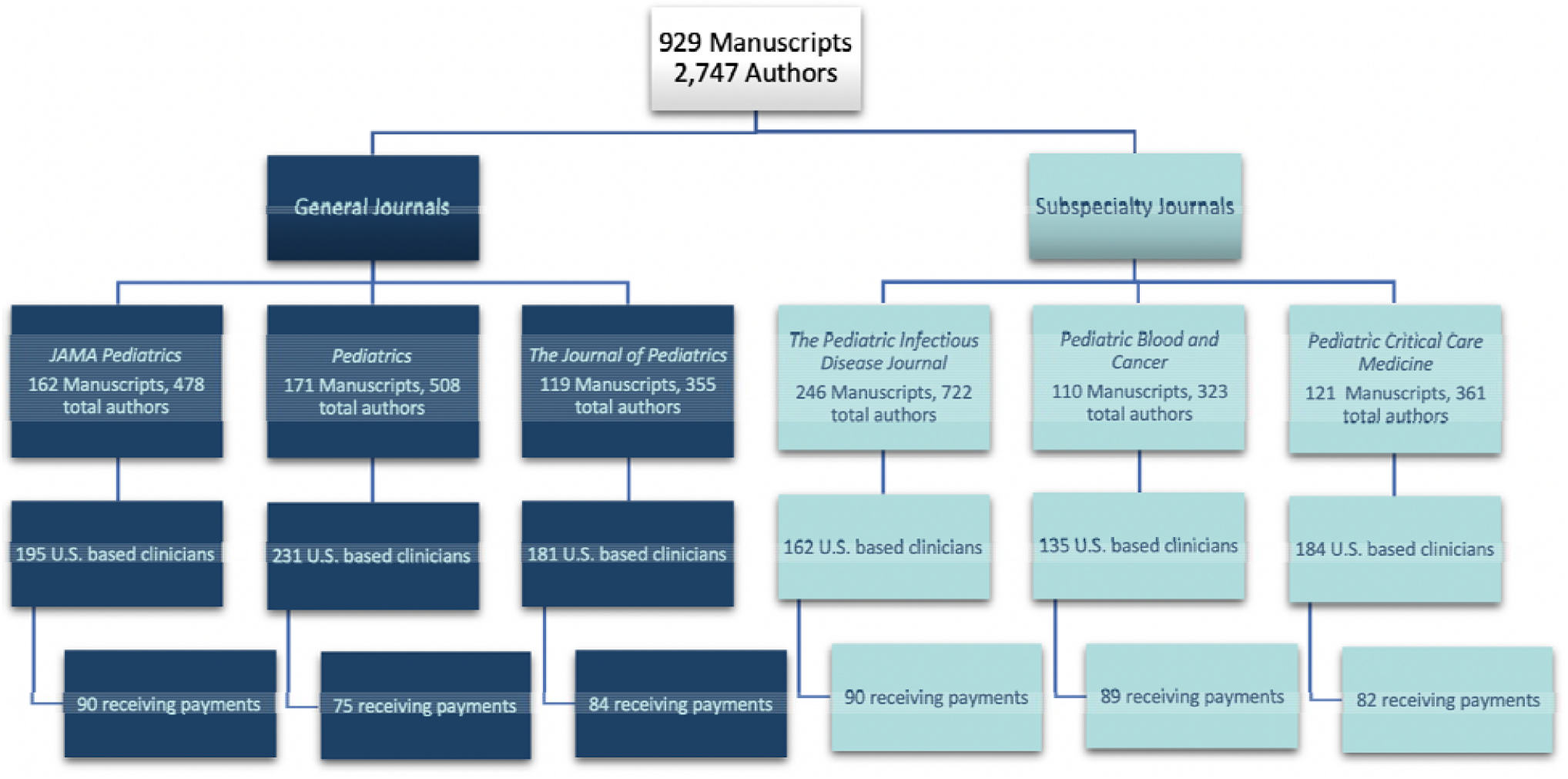
Flow diagram showing the pediatric journals, manuscripts, and authors receiving industry payments as reported by ProPublica’s Dollars for Docs.

There was a strong correlation between data collected from PDD and CMSOP (r=1.00, p < .0001, Supplemental Figure 1), which ensured accuracy of recording of payments.

Two-fifths (41.0%) of authors who had published in general (*JAMA Pediatrics, Pediatrics*, or *The Journal of Pediatrics*) journals had received payments, while over half (54.3 %) of those who had published in subspecialty (*Pediatric Critical Care Medicine, Pediatric Infectious Disease Journal*, or *Pediatric Blood and Cancer*) journals had received payments. Authors who had published in subspecialty journals were 32.3% more likely to have received payments than those who had published in the general pediatrics journals (RR=1.32, 95% CI 1.16-1.50, p<0.0001). Although the mean amount received was higher for authors of articles in the general pediatrics journals (mean difference = 5,640.83, 95% CI = (-6,377.37, 17,659.03) and p-value = 0.357, Supplemental Figure 2), the difference was not statistically significant.

**Figure 2.**
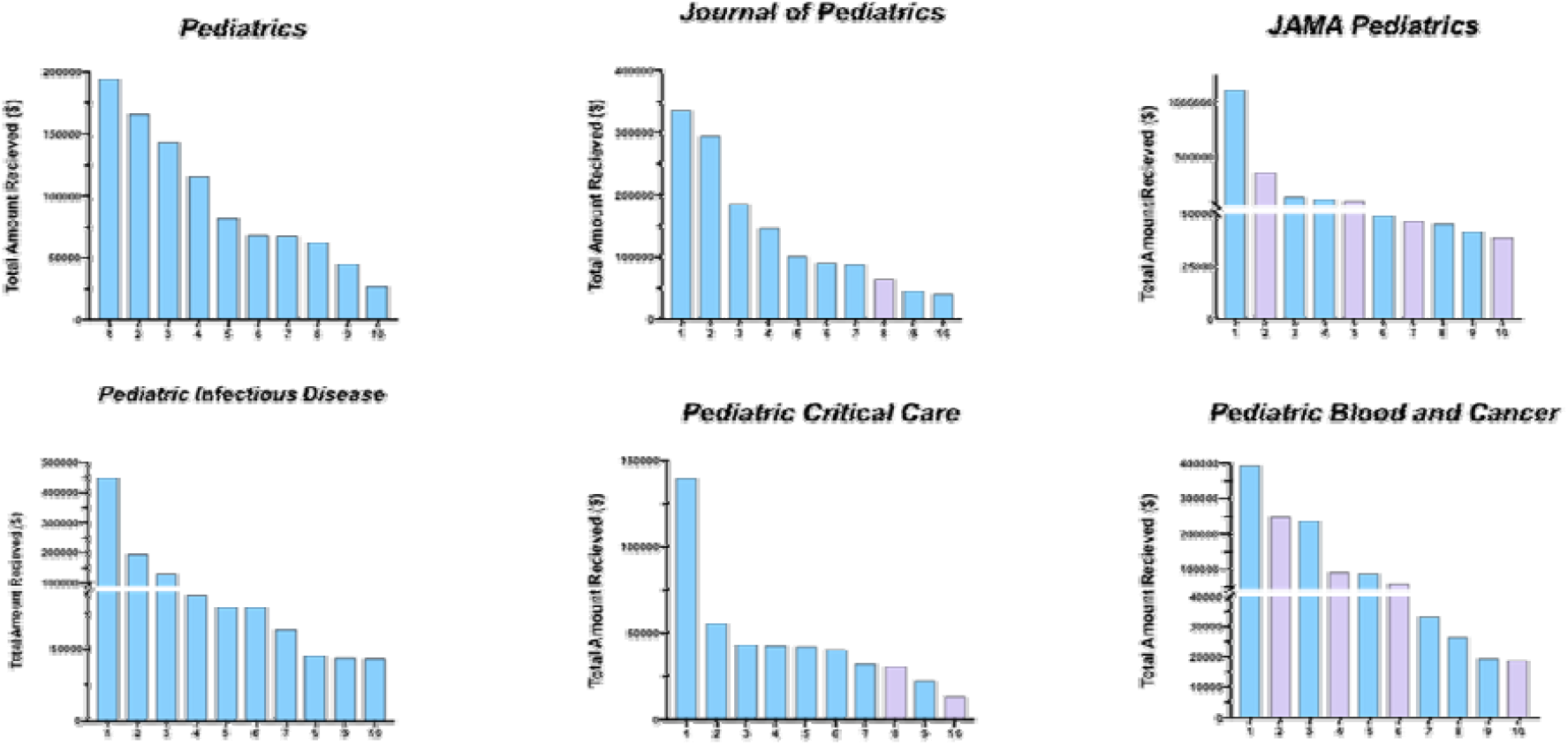
Top ten compensated authors as reported by ProPublica’s Dollars for Docs (2015-2018) for general (top) and sub-specialty (bottom) pediatrics journals. Male authors (49) are shown in blue and females (11) in pink.

Of 438 female authors, 42.2% received payments. Overall, males were 19.6% more likely to have received payments than females (RR=1.20, 95% CI 1.05-1.37, p = 0.008). The mean amount received by males was $23,250.71 while that received by females was $10,970.78 (mean difference = 12,279.924, 95% CI = (2036.31, 22,523.54), p = 0.019, Supplemental Figure 3). The majority of top ten earners from each journal were males (Figure 2). Of 643 male authors who met inclusion criteria, 50.5% received payments.

## Discussion

Over 45% of U.S. based physicians who authored articles published in six high impact pediatric journals had received payments of at least $10 or more within 36 months prior to their publication. Physicians who had published in a subspecialty journal were more likely to have received payments.

Disparities between payments to male and females were also observed. Of the physicians who received payments, males were both significantly more likely to have received payments and, on average, had received higher amounts than females. These results align with previous findings that have documented a disparity between industry payments to male and female physicians of various specialties, including male and female otolaryngologists and radiation oncologists (12, 13). One report that examined payments to practicing physicians of various specialties found that women received approximately $3,600 per year fewer total dollars from industry in comparison to men (14). Because relationships between industry and physicians have the potential to increase understanding of and access to new technology and drugs, the disparity in these relationships between males and females may ultimately have a negative outcome on a female physician’s advancement of her career (12).

Disparities between male and female physicians, beyond industry payments, are well documented. For example, previous analyses have found that on average female physicians are paid less, are less likely to publish articles, speak at conferences, or obtain grant funding (18, 19). These disparities persist despite increasing numbers of females entering medicine, including pediatrics (15). This may be due to lack of access to mentoring, childcare responsibilities, and other non-career-oriented responsibilities (13). Further research is needed to accurately elucidate the cause of these differences in payments to pediatricians.

We found a perfect correspondence between CMSOP and PDD for amount received and the number of payments. A prior investigation noted that, on rare occasions, there were occasionally discrepancies between these databases (8) which presumably was due to contested payments which were corrected in CMSOP before PDD.

There are some important limitations to note for this study. First, the study only considers physicians practicing in the US and thus does not account for the potential CoIs of authors who hold other degrees or who live outside the US, which is a group that accounted for 60.4% of authors. Similar tools have been developed in Australia (19) and the Netherlands (20), but no studies have examined cross country comparisons. Future studies should example CoI among non-US authors and the extent to which these disparities persist across countries. Second, choosing the middle author based on receiving payments may have artificially elevated the values we obtained. However, when we looked at articles rather than authors, we found that 68% of articles had at least one U.S. physician who had received payments

Overall, we determined that two-fifths of pediatric authors in high impact journals had significant CoIs totaling $9.6 million. We also observed sex differences between the payments received, and thus potential CoIs, of male and female pediatric journal authors. Future studies should assess the accuracy of disclosure as well as monitor the trends in the CoIs of journal authors and sex disparities among authorship and industry sponsorship. Because CoIs will continue to have the potential to influence patient welfare and research integrity, future studies will be important in continuing to quantify and describe potential CoIs as new data becomes available annually.

## Data Availability

The data obtained in this study is publicly available.

https://projects.propublica.org/docdollars/

## List of Acronyms

(CoI): Conflict of Interest
(CMSOP): Centers for Medicare and Medicaid Services Open Payments
(PDD): ProPublica’s Dollars for Docs

## Disclosures

BJP is part of an osteoarthritis research team supported by Pfizer and Eli Lilly. The other authors have no disclosures.

## Supplemental Figures

**Supplemental Figure 1.**
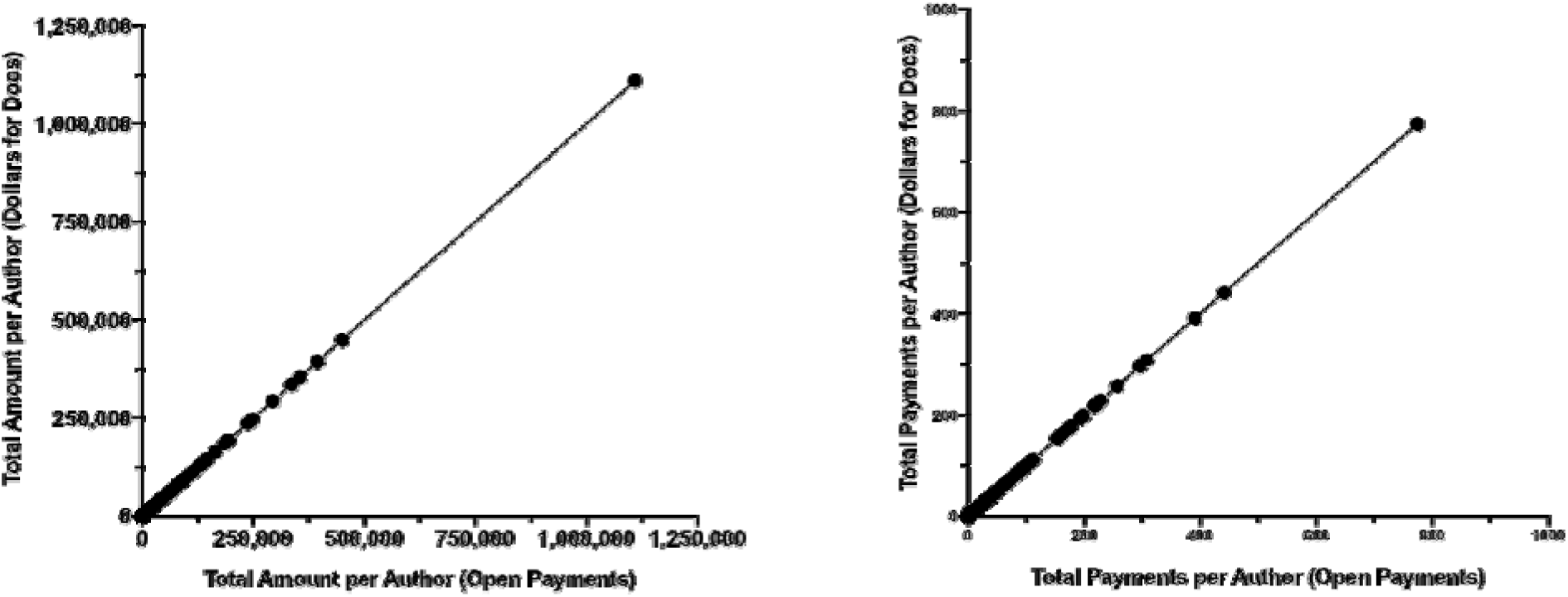
Perfect agreement (r = 1.00) between the amount (left) or total payments (right) received per pediatrician-author between the Center for Medicare and Medicaid Service’s Open Payments and ProPublica’s Dollars for Docs.

**Supplemental Figure 2.**
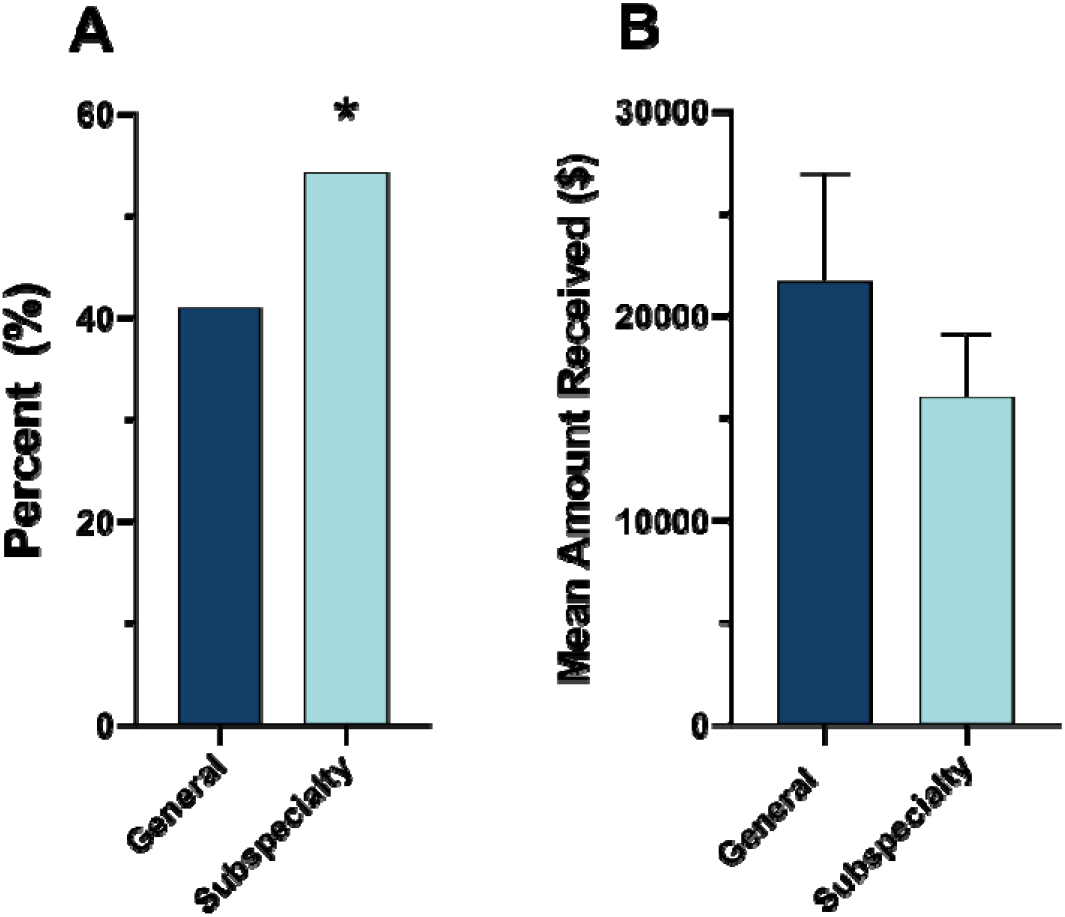
Percent of general (JAMA Pediatrics, Pediatrics, The Journal of Pediatrics) and subspecialty (Pediatric Blood and Cancer, Pediatric Critical Care Medicine, and The Pediatric Infectious Disease Journal) journal author with an entry in ProPublica’s Dollars for Docs (A) and the mean (±SD) received. * p < .05.

**Supplemental Figure 3.**
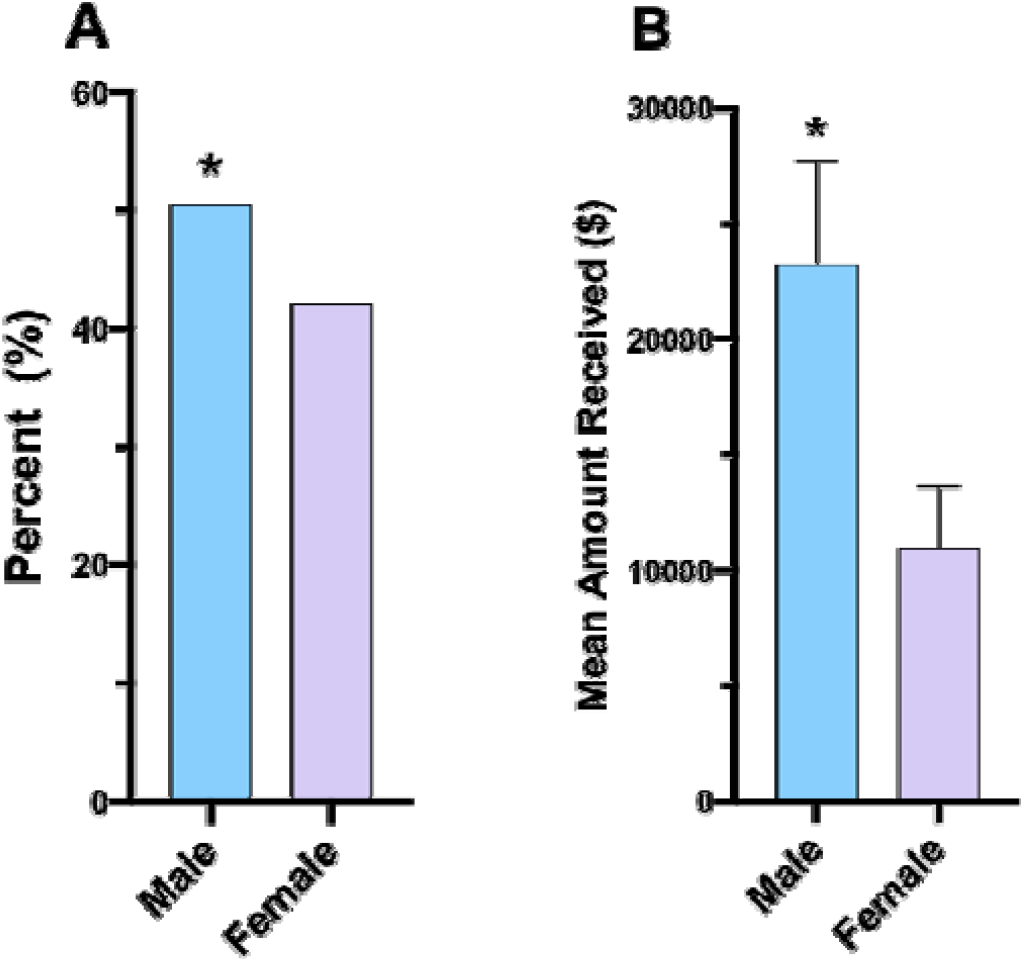
Sex differences in the percent (A) of JAMA Pediatrics, Pediatrics, The Journal of Pediatrics, Pediatric Blood and Cancer, Pediatric Critical Care Medicine, and The Pediatric Infectious Disease Journal) authors with an entry in ProPublica’s Dollars for Docs and the mean (±SD) received. * p < .05.

## Notes

### Funding Statement

No direct external funding was received.

### Author Declarations

Procedures were approved as exempt by the Institutional Review Board of Geisinger.

